# Prospective validation of an uncertainty-aware machine learning model for secondary progressive multiple sclerosis classification

**DOI:** 10.64898/2026.10.07.26364939

**Authors:** Akshai Parakkal Sreenivasan, Yassine Noui, Ida Erngren, Ola Spjuth, Joachim Burman, Kim Kultima

## Abstract

**Background:** Relapsing–remitting multiple sclerosis (RRMS) often progresses to secondary-progressive MS (SPMS), but diagnosis is typically retrospective, delaying potential interventions. Existing rule-based and machine learning approaches often lack prospective validation and provide no uncertainty estimates, limiting clinical adoption.

**Objective:** To prospectively evaluate a conformal prediction (CP)-enabled random forest model for reproducing clinician-recorded disease course labels in the Swedish Multiple Sclerosis Registry (SMSReg) and compare its classifications with the Lorscheider criteria.

**Methods:** The CP model was trained and calibrated on SMSReg-2022 (10,787 individuals, 85,461 hospital visits) and then validated on SMSReg-2025 (15,719 individuals, 45,077 hospital visits; not included in the SMSReg-2022 training/calibration/testing datasets). Performance was assessed at the hospital visit and individual levels and compared with the Lorscheider criteria.

**Results:** The model maintained good calibration on the prospective dataset. At the per-hospital visit-level, the model accuracies were 92% (RRMS) and 95% (SPMS). At the individual level, sensitivity was 98%, specificity 89%, and F1-score 0.75. With clinician-recorded labels as reference, the model showed higher sensitivity than the Lorscheider criteria (98% vs. 71%), more transition assignments (90% vs. 57%), and higher F1-score (0.76 vs. 0.69).

**Conclusion:** The CP-enabled model prospectively reproduced clinician-recorded disease course labels with calibrated uncertainty, supporting harmonization and decision support.

## Introduction

Multiple sclerosis (MS) is among the most common and important causes of neurological disability in the young adult population[1]. While the diagnosis of MS and the possibility of prompt intervention in the earliest stages of relapsing-remitting disease (RRMS) have improved with recent revisions of the McDonald diagnostic criteria [2,3], considerable challenges remain in the timely recognition of the secondary progressive MS (SPMS) disease course. Increasing evidence suggests that MS progression lies on a continuum rather than in sharply defined subtypes [4], which may underlie the retrospective assignment of SPMS. This assignment, unlike the McDonald criteria for RRMS or primary progressive MS (PPMS), does not include any quantitative or ‘objective’ requirements. This, alongside a relative paucity of effective treatments for SPMS [5,6], contributes to delays in the recognition of SPMS, a lag measured at approximately three years in previous studies [7]. More recently, the Bruton’s tyrosine kinase (BTK) inhibitor tolebrutinib has been shown to be effective in preventing disability accumulation in individuals with SPMS with little effect on relapsing disease [8–10], underscoring the need for earlier and more accurate detection of the RRMS-to-SPMS transition.

Several efforts have attempted to address this gap [11–15]. The rule-based method developed by Lorscheider et al. (henceforth referred to as the Lorscheider criteria) is among the most commonly used and well-established approaches, demonstrating good sensitivity and specificity [15]. However, its reliance on longitudinal data limits its applicability to real-world clinical practice. Other methods have been utilized, including a decision tree classifier based on cross-sectional clinical data, which provides predictions at individual hospital visits [14]. Despite these advances, none of these methods provides uncertainty estimates, a critical feature for clinical decision-making when predictions are uncertain [16].

In our previous work, we developed a random forest model trained on data from the Swedish MS registry (SMSReg) [17], integrating conformal prediction (CP) to provide individual-specific uncertainty estimates and explainable artificial intelligence (XAI) methods to enhance interpretability [18]. This model demonstrated strong performance in distinguishing between RRMS and SPMS at each hospital visit when evaluated using SMSReg data. A key limitation, shared by prior work, was the lack of prospective validation, an essential step bridging experimental performance to clinical practice.

In this study, we address this gap by evaluating the Swedish model, trained on SMSReg data up to 2022, using prospectively collected data from 2025 [17]. We benchmarked model classifications against the established Lorscheider criteria to assess agreement, relative strengths, and limitations in a contemporary registry setting.

## Material and methods

### Ethical approval

This study adhered to the Declaration of Helsinki and received approval from the Swedish Ethical Review Authority (Dnr 2021-00702, 2024-07442-02, and 2025-00541-02). The requirement for informed consent was waived, as the research used anonymized registry data from individuals who had previously consented to research use.

### Data

This study utilized two versions of the SMSReg data: SMSReg-2022 and SMSReg-2025 [17]. The SMSReg-2022 dataset was extracted on 19 December 2022 and the SMSReg-2025 dataset on 21 February 2025, providing an additional 2 years and 2 months of follow-up. SMSReg-2022 contained 22,748 people with multiple sclerosis (pwMS) and 197,227 hospital visits, while SMSReg-2025 contained 24,441 pwMS and 228,192 hospital visits. The data were provided without direct identifiers, and the authors did not have access to the re-identification key.

Data cleaning followed the procedure described in the previous work, where records with duplicates or missing essential information, such as the Expanded Disability Status Scale (EDSS) score, date of birth, disease course (RRMS, SPMS) at each hospital visit, or onset date, were removed [18]. The disease course was assessed by the treating clinician for each pwMS and recorded in SMSReg. The dataset included individuals who transitioned from RRMS to SPMS (pwMS who are transitioning, pwMS-T), individuals who remained RRMS throughout follow-up (people with RRMS, pwRRMS), and individuals who remained SPMS throughout follow-up (people with SPMS, pwSPMS).

After quality control and exclusion of people with PPMS, the SMSReg-2022 dataset contained 14,382 individuals and 113,784 hospital visits, while SMSReg-2025 contained 19,004 individuals and 158,722 hospital visits. Of these, 45,077 hospital visits from 15,719 individuals were exclusive to SMSReg-2025 (Table 1). Individuals reclassified from SPMS to PPMS between 2022 and 2025 were excluded from the analysis. Some individuals previously omitted from SMSReg-2022 due to missing baseline data were newly included in SMSReg-2025, likely reflecting an increase in data completeness over time. For pwRRMS, all hospital visits within two years of the last recorded visit were excluded in SMSReg-2022 to minimize the risk of including undetected transitions during model training and evaluation. In contrast, no cutoff was applied for the present study (SMSReg-2025). Consequently, pwRRMS data in SMSReg-2022 extended at most to 2020, although individual-level trimming sometimes limited follow-up to earlier years. Visits from these trimmed periods were therefore included in SMSReg-2025, which comprised both newly registered individuals and those present in SMSReg-2022 but excluded from model development (training, calibration, or testing), explaining the higher number of visits in both SMSReg-2025 and exclusive SMSReg-2025.

**Table 1.** Data splits used in the study.

| Dataset | PwRRMS | PwSPMS | PwMS-T | RRMS visits | SPMS visits |
| --- | --- | --- | --- | --- | --- |
|  | **Training** |  |  |  |  |
| **SMSReg-2022** | 6,905 | 1,821 | 1,341 | 59,898 | 19,792 |
| **SMSReg-2025** | 6,751 | 1,827 | 1,474 | 80,777 | 21,684 |
| **Exclusive SMSReg-2025** | 6,597 | 1,092 | 88 | 20,989 | 1,877 |
|  | **Calibration** |  |  |  |  |
| **SMSReg-2022** | 467 | 152 | 101 | 4,200 | 1,571 |
| **SMSReg-2025** | 458 | 152 | 110 | 5,559 | 1,727 |
| **Exclusive SMSReg-2025** | 443 | 95 | 5 | 1,368 | 154 |
|  | **Testing** |  |  |  |  |
| **SMSReg-2022** | 2,451 | 677 | 467 | 21,388 | 6,935 |
| **SMSReg-2025** | 6,369 | 1,257 | 606 | 39,201 | 9,774 |
| **Exclusive SMSReg-2025** | 6,318 | 956 | 125 | 17,877 | 2,812 |

Disease course (RRMS or SPMS) was taken from the clinician-recorded disease course and used as the reference label. pwRRMS (people with RRMS) were those diagnosed with RRMS at both the initial and final visit; pwSPMS (people with SPMS) had an SPMS diagnosis at both the initial and final visit; and pwMS-T (people with MS who are transitioning) had RRMS at the initial visit and SPMS at the final visit. The SMSReg-2022 dataset was used for model training, calibration, and testing in the previous work[18]. For SMSReg-2025 and exclusive SMSReg-2025, the table shows the corresponding individuals matching the SMSReg-2022 data splits. Individuals not present in SMSReg-2022 were included in the SMSReg-2025 test set. SMSReg-2025 contains all individuals in the dataset, whereas exclusive SMSReg-2025 includes only hospital visits unique to SMSReg-2025.

Variables were harmonized with those used in the previous study, resulting in a total of 27 features [18]. Each hospital visit included the individual’s age and EDSS score at the time of the visit. These data were supplemented with the most recent available clinical measures: treatment history, results from clinical assessment tests, relapse data, and MRI findings, along with the age at which these measures were recorded. Treatments were classified into first-line or second-line disease-modifying therapies, relapse treatments, stem cell therapies, and other drugs. Relapse-related variables captured the cumulative number of specific relapse types (unilateral optic neuritis, sensory/afferent monofocal relapse, multifocal relapse, and steroid-treated relapse) up to the visit date, as well as binary indicators for treatment and remission of the most recent relapse. MRI data included counts of T2-weighted lesions and the number and site of T1-weighted gadolinium-enhancing lesions (brain or spinal cord), with lesion counts categorized as ≤ 9, > 9 and ≤ 20, or > 20.

### Model

The random forest model used in this study, which integrates CP for uncertainty quantification [19] and SHapley Additive exPlanations (SHAP) for interpretability [20], was developed in the previous work [18]. The analyses were performed using Python 3.9. The random forest model was implemented using scikit-learn 1.6.1, and SHAP values were calculated using shap 0.48.0. The model was configured with 150 estimators, a minimum of five samples per leaf, the Gini impurity criterion, balanced class weights, and no maximum depth restriction. The model was trained on SMSReg-2022 data comprising 10,067 pwMS and 79,690 hospital visits, and calibrated on a subset of 720 pwMS with 5,771 hospital visits. The code used for the analysis is available at https://github.com/caramba-uu/MSP-tracker_prospective_analysis.git

### Conformal prediction

To incorporate uncertainty quantification into the predictions, we employed conformal prediction (CP), a model-agnostic framework for assessing predictive uncertainty [19]. Specifically, we used inductive CP, which calibrates model outputs on a separate calibration set. CP was implemented using nonconformist 2.1.0.

At a significance level of alpha, CP predictions can be single-label ({RRMS} or {SPMS}), multi-label ({RRMS, SPMS}), or empty ({}). Single-label predictions are considered confident because they satisfy the user-specified significance level and are therefore preferred. Multi-label predictions, although not incorrect, reflect cases in which the model is uncertain about assigning a unique class at the chosen significance level. Such predictions may signal the need for further expert evaluation. Empty predictions represent instances in which the model refrains from providing an output, typically due to distributional shifts between the calibration and test data.

A central property of CP is validity, which ensures that the empirical error rate matches the expected error rate over time. This can be evaluated by examining calibration quality, typically by plotting the expected versus observed error rates (calibration plot). A well-calibrated model yields a curve close to the diagonal, whereas deviations indicate miscalibration. To assess whether calibration was maintained, the model was calibrated on the SMSReg-2022 dataset, and its validity was subsequently evaluated on data exclusive to the SMSReg-2025 dataset.

### Lorscheider criteria

The Lorscheider criteria was applied as an established rule-based comparator for SPMS classification[15]. According to the criteria, SPMS onset is defined as disability progression confirmed over at least three months in the absence of relapse activity. Progression is indicated by an EDSS increase of 1.0 when baseline EDSS ≤5.5 or 0.5 when ≥ 6.0. Additionally, the EDSS must reach a minimum of 4.0 and be accompanied by a pyramidal functional system (FS) score of 2 or higher. FS scores were incorporated into the analysis whenever available for a given hospital visit.

### Statistical analysis

To assess changes in model misclassification over follow-up in pwRRMS, the analysis was limited to the last five years of follow-up, measured backward from each individual’s most recent visit. The misclassification rate was calculated as the proportion of visits misclassified by the model within each one-year interval and is presented with 95% Wilson confidence intervals. Misclassification rates between intervals were compared using one-sided two-proportion z-tests to assess whether misclassification was higher in more recent intervals than in earlier intervals. All pairwise comparisons among the five intervals were performed, resulting in 10 comparisons. A Bonferroni correction was applied to account for multiple comparisons. Statistical significance was defined as an adjusted p-value < 0.05. Statistical analyses were performed using statsmodels 0.14.5.

## Results

The study utilized two versions of the SMSReg dataset, SMSReg-2022 and SMSReg-2025, each containing clinical data for pwMS and their hospital visits. After quality control and data splitting, baseline characteristics of the study population are shown in Table 2. The exclusive SMSReg-2025 dataset, containing hospital visits unique to SMSReg-2025, was combined into a single test set for evaluation in this study. Model performance was assessed at a CP significance level of 0.07 (confidence of 93%), which had previously been identified as optimal for maximizing single-label predictions in SMSReg-2022 [18].

**Table 2.**
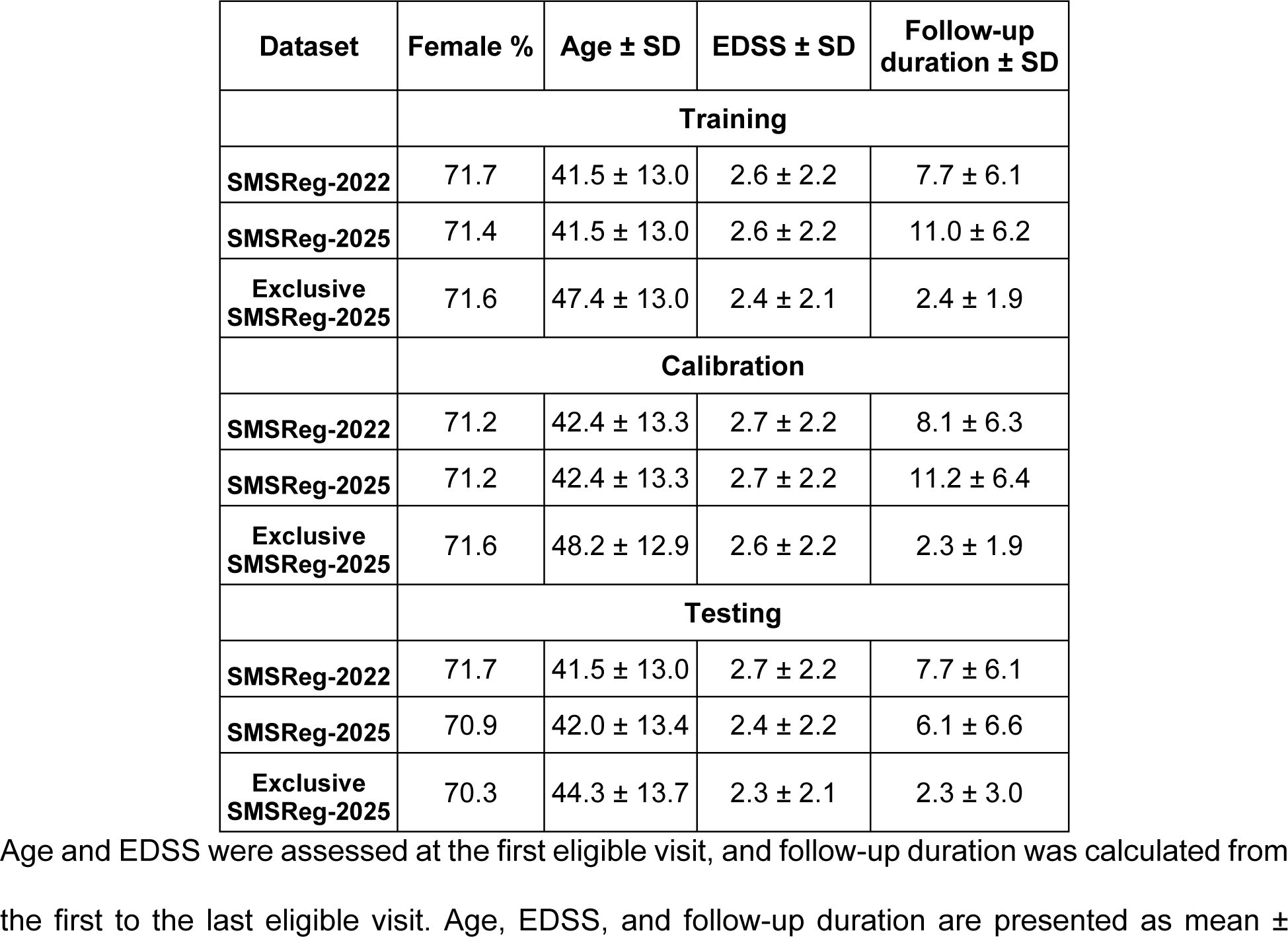
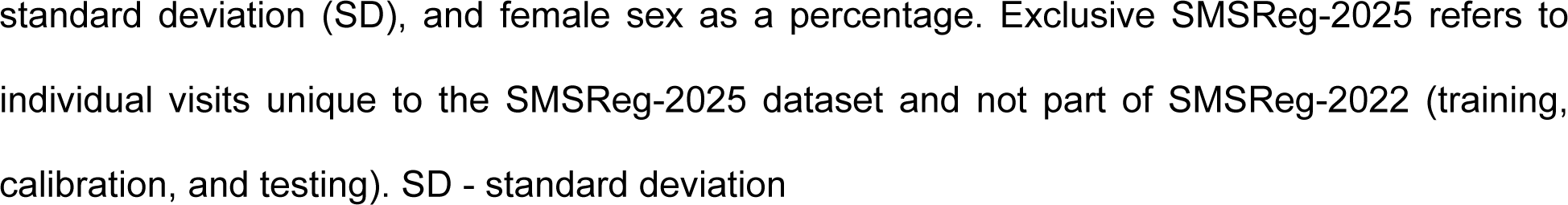
Baseline characteristics of individuals in each dataset.

| Dataset | Female % | Age ± SD | EDSS ± SD | Follow-up duration ± SD |
| --- | --- | --- | --- | --- |
| **Training** |  |  |  |  |
| **SMSReg-2022** | 71.7 | 41.5 ± 13.0 | 2.6 ± 2.2 | 7.7 ± 6.1 |
| **SMSReg-2025** | 71.4 | 41.5 ± 13.0 | 2.6 ± 2.2 | 11.0 ± 6.2 |
| **Exclusive SMSReg-2025** | 71.6 | 47.4 ± 13.0 | 2.4 ± 2.1 | 2.4 ± 1.9 |
| **Calibration** |  |  |  |  |
| **SMSReg-2022** | 71.2 | 42.4 ± 13.3 | 2.7 ± 2.2 | 8.1 ± 6.3 |
| **SMSReg-2025** | 71.2 | 42.4 ± 13.3 | 2.7 ± 2.2 | 11.2 ± 6.4 |
| **Exclusive SMSReg-2025** | 71.6 | 48.2 ± 12.9 | 2.6 ± 2.2 | 2.3 ± 1.9 |
| **Testing** |  |  |  |  |
| **SMSReg-2022** | 71.7 | 41.5 ± 13.0 | 2.7 ± 2.2 | 7.7 ± 6.1 |
| **SMSReg-2025** | 70.9 | 42.0 ± 13.4 | 2.4 ± 2.2 | 6.1 ± 6.6 |
| **Exclusive SMSReg-2025** | 70.3 | 44.3 ± 13.7 | 2.3 ± 2.1 | 2.3 ± 3.0 |

Age and EDSS were assessed at the first eligible visit, and follow-up duration was calculated from the first to the last eligible visit. Age, EDSS, and follow-up duration are presented as mean ±
standard deviation (SD), and female sex as a percentage. Exclusive SMSReg-2025 refers to individual visits unique to the SMSReg-2025 dataset and not part of SMSReg-2022 (training, calibration, and testing). SD - standard deviation

### Model performance

Calibration curves demonstrated that the model remained well-calibrated on this prospective dataset, with observed error rates closely matching expected error rates (Fig. 1A). Considering all hospital visits, the model produced correct single-label predictions for 92% (36,917/40,234) of RRMS visits and 95% (4,588/4,843) of SPMS visits (Fig. 1B). The model produced 45,012 (99.9%) single-label predictions, 65 (0.1%) two-label prediction sets, and no empty sets.

**Fig. 1.**
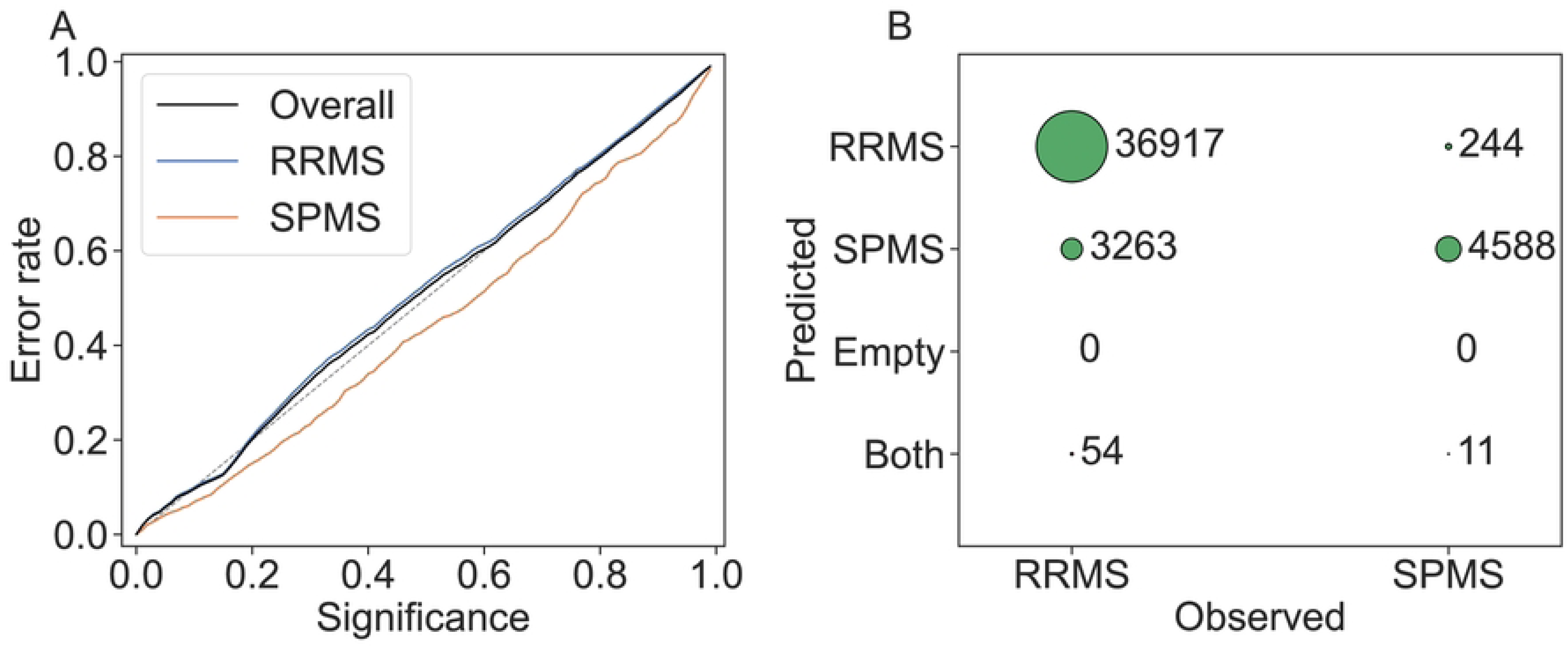
Overall performance on the test set. A) Calibration curve showing the expected error rate (significance) on the x-axis and the observed error rate on the y-axis. The diagonal lines for RRMS, SPMS, and overall indicate a well-calibrated model for the test set. B) Confusion matrix showing predictions at a significance level of 0.07 (93% confidence) for all the hospital visits.

We further assessed the model’s ability to classify on a per-individual basis. Individuals were labeled as SPMS if the model predicted SPMS at any visit, and as RRMS otherwise. The test set comprised 13,358 pwRRMS, 2,143 pwSPMS, and 218 pwMS-T, corresponding to an overall SPMS proportion of 15%. The model achieved a sensitivity of 98%, a specificity of 89%, a positive predictive value (PPV) of 61%, and a negative predictive value (NPV) of ∼100%, with an F1-score of 0.75 (Fig. 2A).

**Fig. 2.**
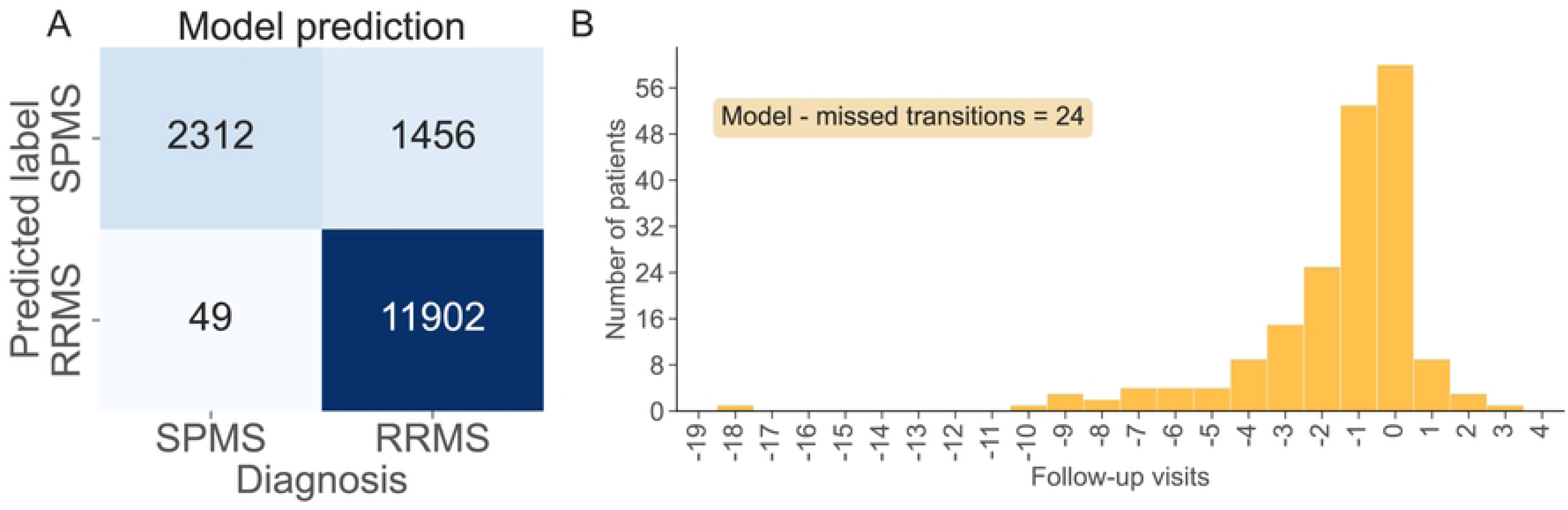
Performance of the model on the entire test set. A) Confusion matrix showing the model’s ability to classify individuals as RRMS or SPMS. Individuals were labeled SPMS if at least one visit was predicted as SPMS and otherwise they were labeled RRMS. B) Time of transition predictions for 218 pwMS-T compared with clinician diagnosis. Negative values indicate that the model predicted the transition earlier than the clinician’s diagnosis, positive values indicate a later prediction, and zero represents predictions made at the same visit as the clinician’s diagnosis. There were 24 missed transitions, indicating cases where the model did not identify the transition to SPMS.

To further analyze model behavior and better understand the lower PPV observed, we evaluated misclassification rates in pwRRMS (S1 Fig.). Significantly higher misclassification rates were observed during the recent follow-up intervals (0–1 and 1–2 years) compared with prior follow-up years. The increased misclassification toward the most recent follow-ups may partly reflect diagnostic delays in identifying SPMS conversion, in which some individuals may have already transitioned but remain clinically recorded as RRMS. However, this possibility cannot be verified within the present study design. This label-timing uncertainty may also contribute to the model’s lower PPV.

Finally, we examined the timing of model-assigned SPMS labels relative to clinician-recorded transition visits (Fig. 2B). Of the 218 pwMS-T, the model predicted transitions in 194 (89%), with 190 (87%) identified at either the exact visit, within one visit, or earlier.

### Model interpretability

To ensure model interpretability, feature importance was assessed using SHAP values (S2 Fig.). EDSS score and age at visit were the most influential features, consistent with previous findings [18], with only minor variations in this dataset. Specifically, EuroQol 5 Dimensions (EQ-5D) and age at Symbol Digit Modalities Test (SDMT) showed slightly greater importance, whereas SDMT score contributed less. These findings demonstrate that the model’s decision patterns remained stable and clinically interpretable when applied to newly acquired registry data.

### Comparison with the Lorscheider criteria

To benchmark the model’s performance, we compared its classifications with those obtained using the Lorscheider criteria. For individuals in the exclusive SMSReg-2025 dataset, those with fewer than three visits in SMSReg-2025 were excluded, as the Lorscheider criteria require longitudinal information, including a baseline EDSS score, evidence of progression, and subsequent confirmation. After applying this criterion, the exclusive SMSReg-2025 dataset consisted of 10,750 pwRRMS, 1,809 pwSPMS, and 202 pwMS-T, corresponding to an SPMS proportion of 15.8%. Given the longitudinal nature of the Lorscheider criteria, earlier visits available for these individuals in the SMSReg-2025 dataset were also incorporated only when applying the criteria to ensure a fair comparison.

Both the Lorscheider criteria and the model were evaluated for their ability to classify individuals as SPMS or RRMS, using clinician-recorded disease course as the reference. Consistent with the model definition, if the criteria assigned an SPMS label at any visit, the individual was classified as SPMS; otherwise, they were classified as RRMS. The model achieved a sensitivity of 98%, specificity of 89%, PPV of 62%, NPV of ∼100%, and an F1-score of 0.76 (Fig. 3A). The Lorscheider criteria yielded a sensitivity of 71%, specificity of 94%, PPV of 68%, NPV of 95%, and an F1-score of 0.69 (Fig. 3B).

**Fig. 3.**
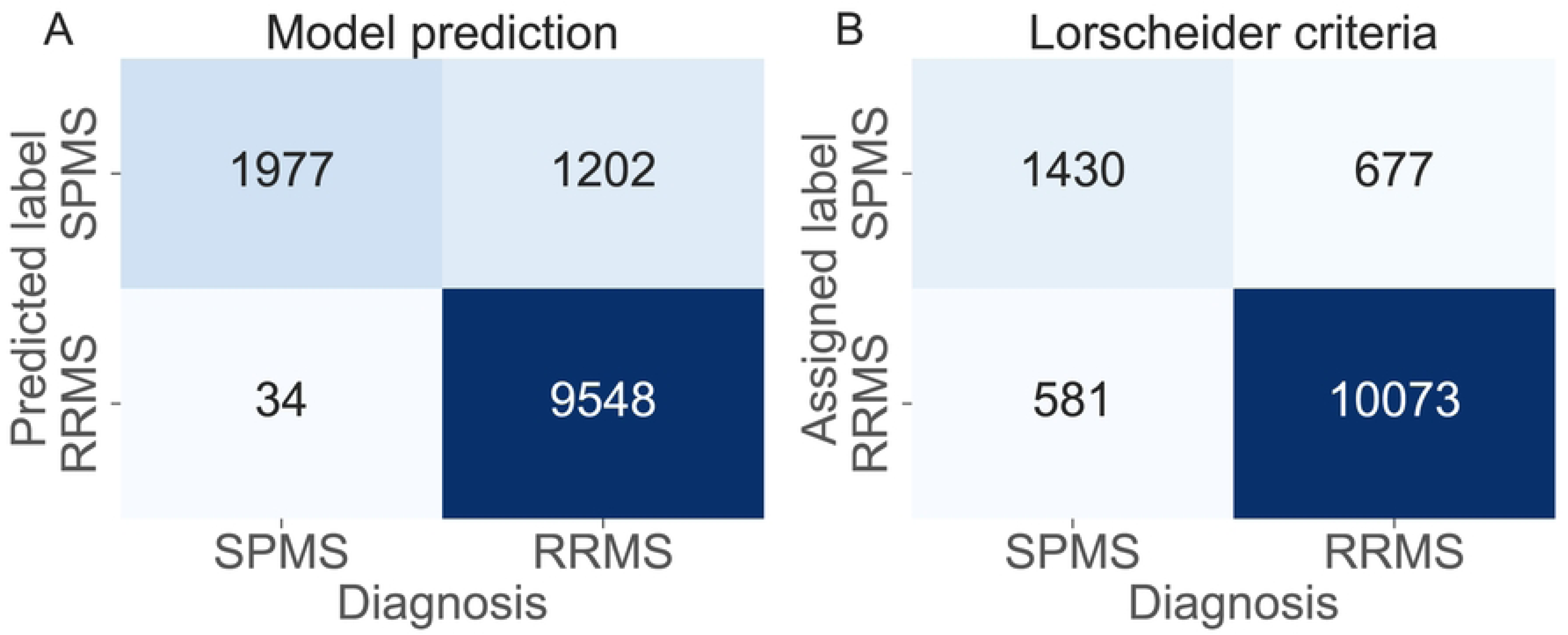
Confusion matrix showing the ability to classify individuals as RRMS or SPMS. Individuals were classified as SPMS if either the model or the criteria assigned an SPMS label at any visit; otherwise, they were classified as RRMS. A) Model. B) The Lorscheider criteria.

To further assess the agreement between the two approaches, we compared their classifications with the clinical diagnosis (Fig. 4). The Lorscheider criteria classified 10,654 individuals as RRMS and 2,107 as SPMS. The model agreed with these classifications in 9,384 RRMS and 1,909 SPMS cases (11,293 of 12,761; 88%), demonstrating strong concordance despite relying solely on cross-sectional data.

**Fig. 4.**
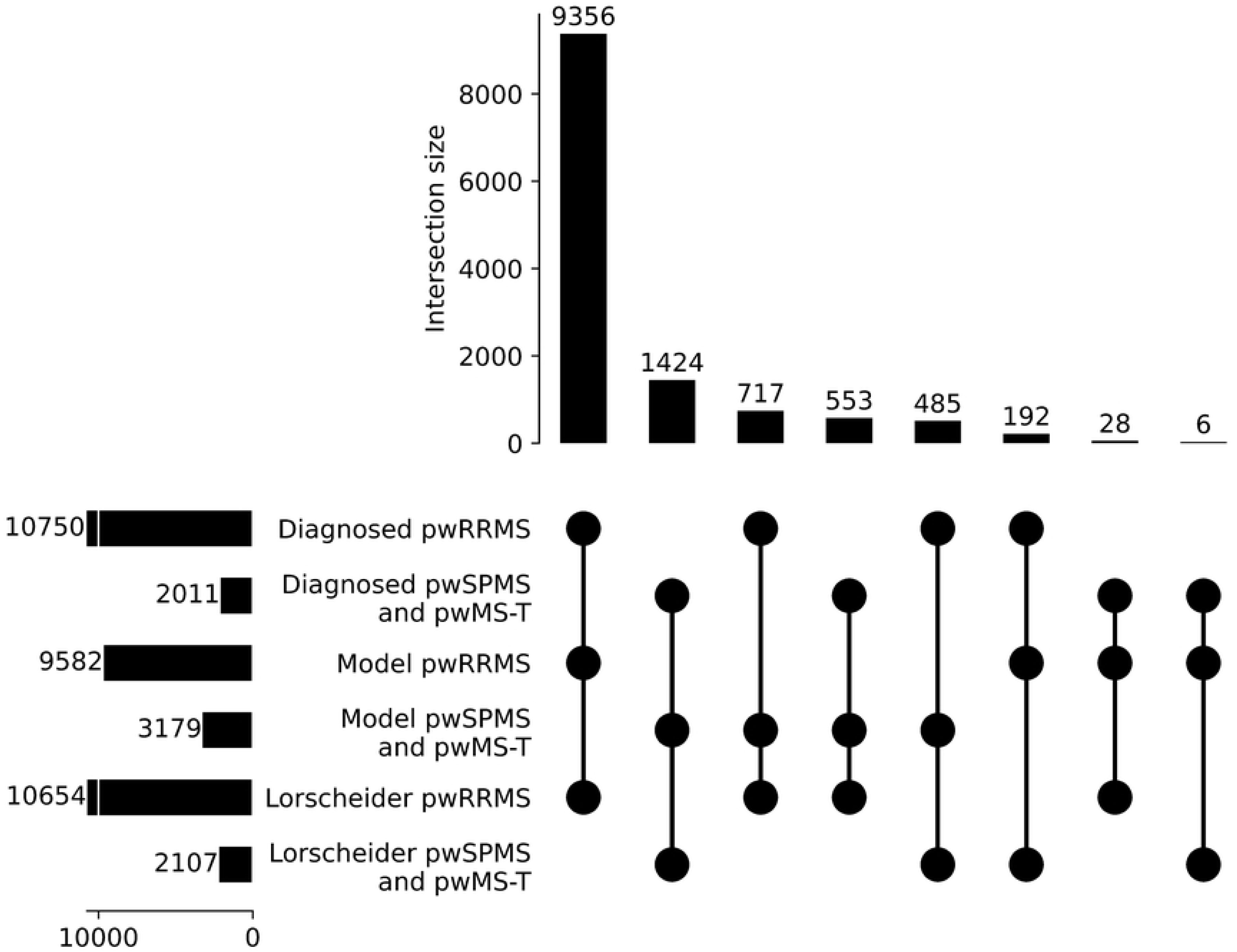
The overlap between clinical diagnosis, model predictions, and Lorscheider criteria classifications of individuals with RRMS (pwRRMS) and SPMS (pwSPMS and pwMS-T). The numbers at the top indicate the degree of overlap between methods. For the 10,654 RRMS according to the Lorscheider criteria, the model predicted RRMS in 9,384 (9,356 + 28) cases, including 28 clinically diagnosed as SPMS. Conversely, 1,270 (717 + 553) were predicted as SPMS, with 553 diagnosed as SPMS and 717 as RRMS. Among the 2,107 individuals classified as SPMS by the Lorscheider criteria, the model predicted 1,909 (1,424 + 485) as SPMS, where 485 were clinically diagnosed as RRMS. The remaining 198 (192 + 6) were predicted as RRMS, with 192 diagnosed as RRMS and 6 as SPMS.

### Comparison with the Lorscheider criteria for identifying transition to SPMS

We next compared the timing of transition assignments between the model and the Lorscheider criteria in the 202 pwMS-T (Fig. 5). The model assigned SPMS at any visit in 181 of 202 pwMS-T (90%), compared with 115 (57%) by the Lorscheider criteria. Notably, the model assigned SPMS at the clinician-recorded SPMS transition visit, within one visit, or earlier in 177 (88%) individuals, compared with 111 (55%) using the Lorscheider criteria.

**Fig. 5.**
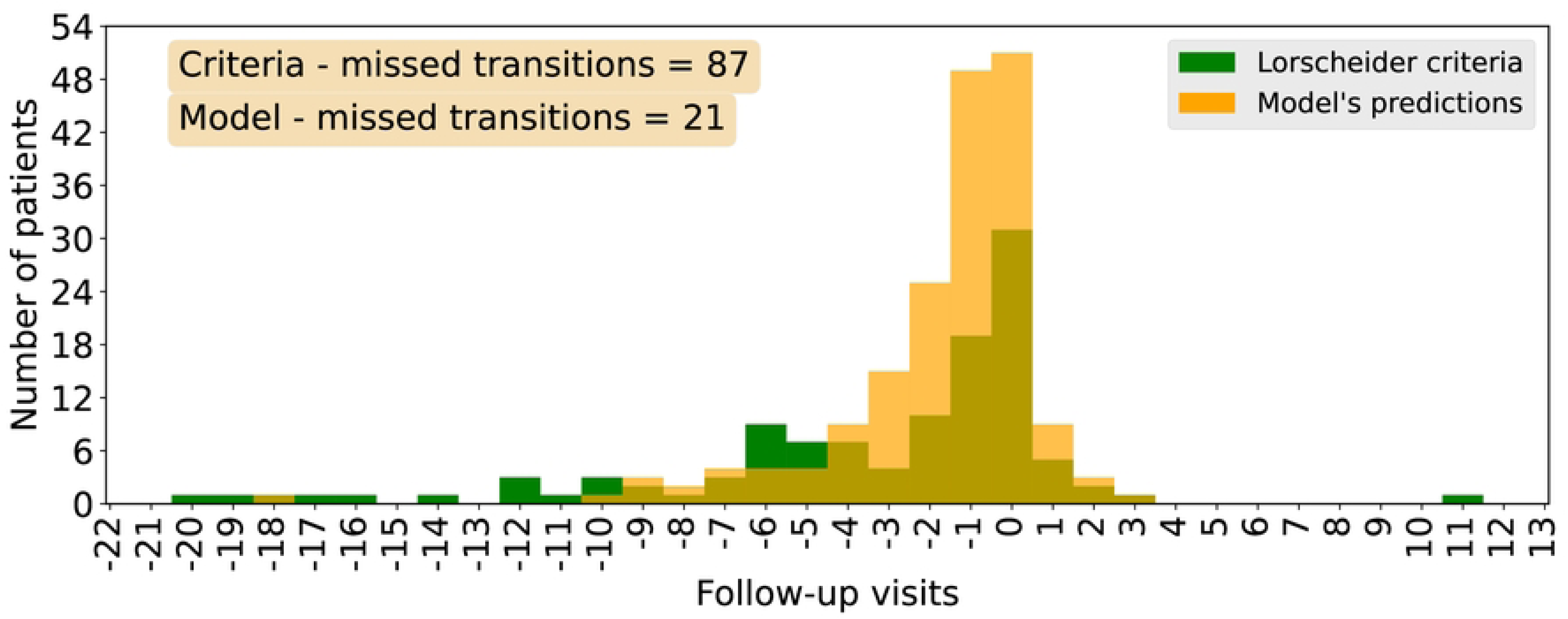
Timing of transition identification for pwMS-T. Model predictions are shown in orange and the Lorscheider criteria in green. The model identified 51 individuals (25%) at the exact visit of transition, 58 (29%) within one visit, 68 (34%) earlier, and 4 (2%) later. In contrast, the Lorscheider criteria identified 31 individuals (15%) at the exact visit, 24 (12%) within one visit, 56 (28%) earlier, and 4 (2%) later. No SPMS assignment was made for 21 individuals by the model and 87 by the Lorscheider criteria. Negative values indicate assignment before the clinician-recorded transition visit, positive values indicate assignment after it, and zero indicates assignment at the same visit.

## Discussion

In this study, we prospectively validated a random forest model augmented with conformal prediction (CP). During the training phase, the model was trained and calibrated using the SMSReg-2022 data [18]. In the validation phase, it was prospectively evaluated on SMSReg-2025, demonstrating notable performance in differentiating SPMS from RRMS at both the hospital visit and individual levels. The model also showed strong temporal agreement, with most model-assigned SPMS labels occurring at or near clinician-recorded transition visits. Importantly, the model delivered robust classifications and provided calibrated uncertainty estimates for each prediction.

These findings extend prior work that has underscored the inherent limitations of retrospective SPMS diagnosis and further highlight the utility of a cross-sectional, interpretable approach suitable for deployment in real-time clinical practice. Moreover, the data set used for evaluation included individuals present in SMSReg-2022 who were not used for model development. It also incorporated two additional years of real-world registry data, thereby providing a stringent and unbiased test set. Such prospective validation is essential for real-life use, as it demonstrates that the model retains accuracy and reliability when applied to new pwMS and evolving registry conditions, closely mirroring its intended application in clinical practice.

### Comparison with the Lorscheider criteria

The Lorscheider criteria remain the most widely used objective standard for SPMS identification, with reported sensitivities of 89% and specificities of 86% in the MSBase dataset [15]. Applied to Swedish registry data, the model demonstrated a 27% increase in sensitivity, an improvement in 5% NPV, and a 0.07-point increase in F1-score compared to the criteria, indicating closer alignment with registry labels. The criteria, on the other hand, showed +5% specificity and +6% PPV, reflecting a more conservative approach. Overall, the model captured more clinician-recorded SPMS cases while maintaining acceptable specificity and improved overall performance.

Among the 202 pwMS-T, the model identified 90% of transitions compared with 57% detected by the Lorscheider criteria. Most model assignments occurred at or near the clinician-recorded SPMS transition visit. These findings indicate closer agreement with clinician-recorded disease course than the Lorscheider criteria, although this may partly reflect the model having been trained on clinician-recorded SMSReg labels. At the same time, the model provides classifications at individual visits, whereas the Lorscheider criteria require longitudinal confirmation of progression.

Other digital tools, such as MSProDiscuss, have also sought to facilitate earlier identification of SPMS by structuring clinical encounters and incorporating patient-reported outcomes [21]. While these tools have demonstrated strong discriminatory ability across RRMS, transitioning, and SPMS states, our model offers a vital complement. It achieves comparable predictive performance using routinely collected registry data and also generates hospital visit-level uncertainty estimates.

A major strength of the current approach is the integration of CP and XAI, which together provide uncertainty quantification and interpretability. Calibration analysis confirmed that the model remained well-calibrated on prospectively collected data, with observed error rates aligning with prespecified thresholds. While not explored in this work, adjusting the significance level would allow for reduced prediction errors at the cost of more multi-label outputs [18,22]. Such outputs can be clinically useful, for instance, to identify individuals at higher risk of transition to SPMS who may warrant closer monitoring. The combination of CP-driven uncertainty quantification and XAI-based interpretability provides clinicians with greater transparency in the model’s outputs; features largely absent from prior approaches but essential for clinical adoption [16]. Together, these features make the approach practical and may help its trustworthiness in a clinical setting.

### Clinical relevance

The importance of timely recognition of SPMS continues to grow as the therapeutic landscape expands. Traditional disease-modifying therapies provide limited benefit once individuals transition to the progressive stage, and may be associated with a higher risk of harm [6,23,24]. Importantly, the recent EU authorization of the BTK inhibitor tolebrutinib for adults with SPMS without relapses in the preceding two years provides a new treatment option specifically targeting this population [8–10]. Early and accurate detection of the RRMS-to-SPMS transition is therefore essential for ensuring that individuals receive the therapy best suited to their disease course. By flagging visits that resemble clinician-recorded SPMS with calibrated uncertainty, the model may prompt timely review and support consistent treatment discussions.

### Limitations and future directions

Alongside the strengths of this study, several limitations warrant consideration. A central limitation is that the reference standard was clinician-recorded disease course rather than independently adjudicated biological transition. Variation in classification practices between neurologists or centres may affect both training labels and measured model performance. The generalizability of the model beyond Sweden remains to be established, as registries and healthcare systems differ in patient populations, documentation practices, and treatment availability. External validation in other registries is therefore essential before broader implementation. Although the reliance on registry data enabled large-scale evaluation, it also highlights opportunities to incorporate additional data types that are not routinely recorded, such as emerging biomarkers (e.g., serum neurofilament light chain), advanced imaging modalities, and glial activation markers. These complementary data sources could further improve early SPMS detection, and future studies could explore integrating them to enhance predictive power. Another limitation is the potential data drift as clinical practices and treatment strategies evolve. Here, CP offers a distinct advantage, as recalibration can preserve validity without requiring full model retraining. The timing of recalibration can be guided by martingale-based methods, which detect distributional drift. Combining martingale monitoring with online CP thus provides a principled framework for maintaining performance over time [25,26]. Finally, prospective clinical implementation studies will be essential to evaluate the real-world impact of the model on clinician decision-making, therapeutic timing, and patient outcomes. These studies should also clarify how best to present uncertainty in ways that support, rather than complicate, clinical care.

## Data Availability

Due to patient privacy regulations, the data underlying the findings of this study cannot be made publicly available. Data may be requested from the Swedish Neuro Registry through a formal data-access application at https://neuroreg.se/forskning/datauttag-for-forskningsandamal/. Data-access applications should be submitted to

https://github.com/caramba-uu/MSP-tracker_prospective_analysis.git

https://neuroreg.se/forskning/datauttag-for-forskningsandamal/

## Acknowledgements

We acknowledge the Swedish Neuro Registries/Multiple Sclerosis Registry for providing data for this study.

## Supporting information

**S1 Fig.**
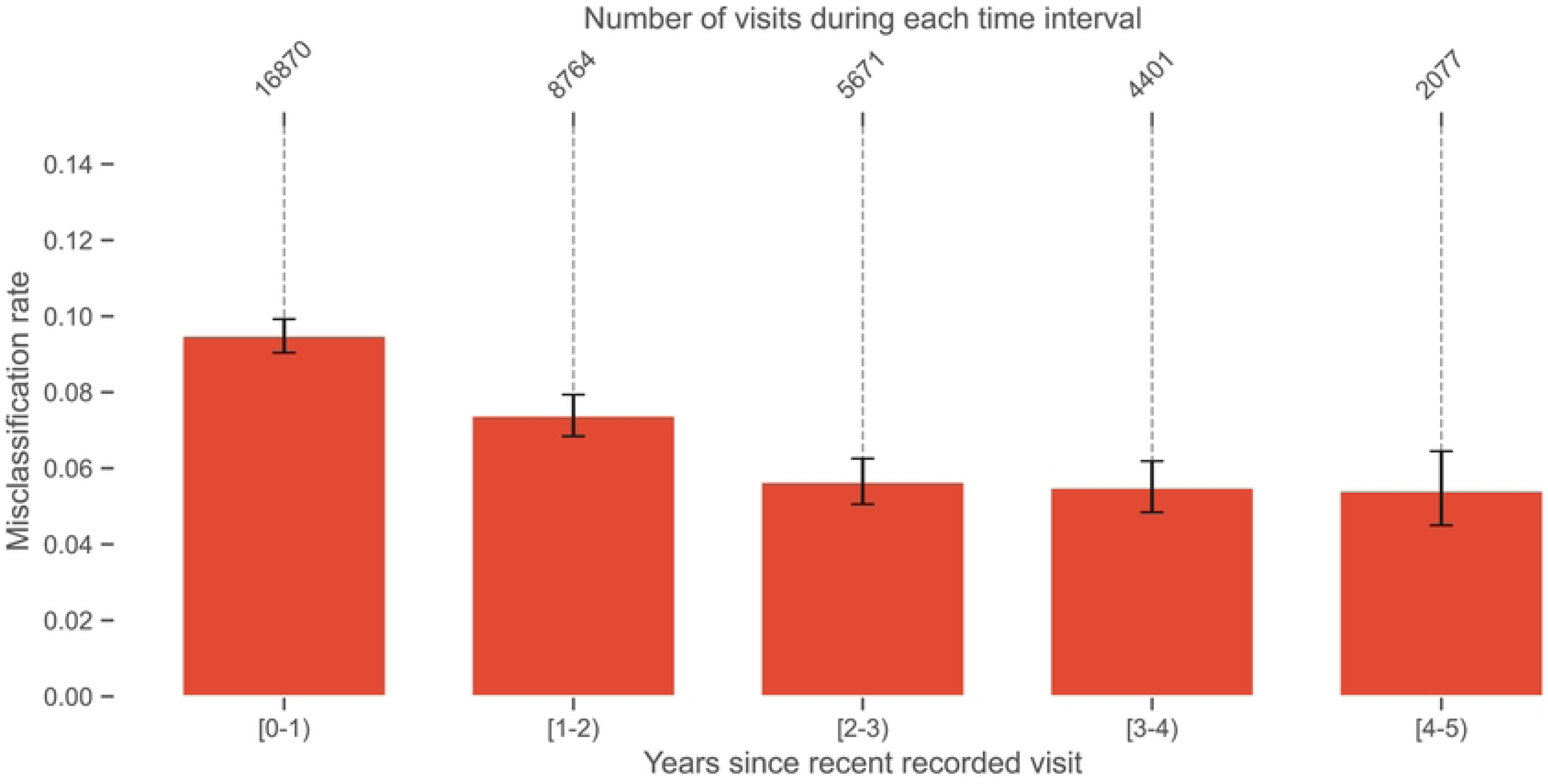
Trend in misclassification of pwRRMS for the model across follow-up years. All pwRRMS were included in this analysis, limited to their last five years of follow-up. The x-axis represents disease duration in years, measured backward from each individual’s most recent visit. The y-axis shows the yearly misclassification rate. Numerical values above the bars indicate the total number of visits within each duration bin. Error bars represent 95% Wilson confidence intervals for each proportion. Visits near the recent follow-up (0–1 and 1–2 years) show significantly higher misclassification rates compared with earlier periods (one-sided two-proportion z-tests, Bonferroni-adjusted for 10 tests, *p* < 0.001). This pattern may partly reflect diagnostic delays in recognizing SPMS conversion, whereby some individuals may already have transitioned but remain clinically recorded as RRMS. However, this possibility cannot be verified within the present study design.

**S2 Fig.**
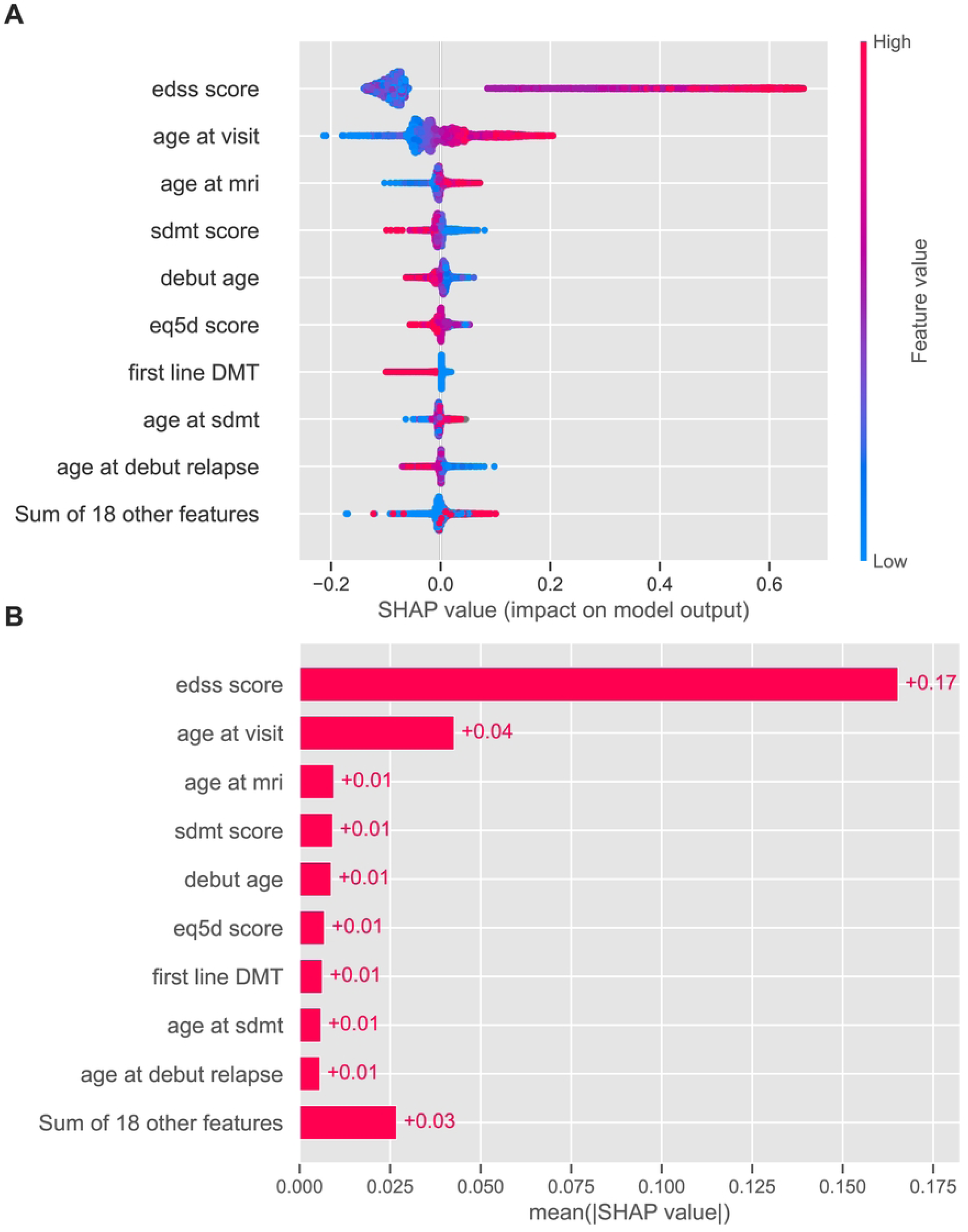
Global feature importance for the test dataset derived from SHAP analysis. (A) Bee swarm plot illustrating the distribution of SHAP values for individual features; positive SHAP values indicate predictions toward SPMS and negative values toward RRMS. Feature values are color-coded, with blue representing lower and red representing higher values. (B) Summary plot showing the mean absolute SHAP values for each feature.

